# Fast and Trustworthy Nowcasting of Dengue Fever: A Case Study Using Attention-Based Probabilistic Neural Networks in São Paulo, Brazil

**DOI:** 10.1101/2025.05.05.25326971

**Authors:** Silas Koemen, Nuno R. Faria, Leonardo S. Bastos, Oliver Ratmann, André Victor Ribeiro Amaral, the Machine Learning & Global Health Network

## Abstract

Nowcasting methods are crucial in infectious disease surveillance, as reporting delays often lead to underestimation of recent incidence and can impair timely public health decision-making. Accurate real-time estimates of case counts are essential for resource allocation, policy responses, and communication with the public. In this paper, we propose a novel probabilistic neural network (PNN) architecture, named NowcastPNN, to estimate occurred-but-not-yet-reported cases of infectious diseases, demonstrated here using dengue fever incidence in São Paulo, Brazil. The proposed model combines statistical modelling of the true number of cases, assuming a Negative Binomial (NB) distribution, with recent advances in machine learning and deep learning, such as the attention mechanism. Uncertainty intervals are obtained by sampling from the predicted NB distribution and using Monte Carlo (MC) Dropout. Using proper scoring rules for the prediction intervals, NowcastPNN achieves nearly a 30% reduction in losses compared to the second-best model among other state-of-the-art approaches. While our model requires a large training dataset (equivalent to two to four years of incidence counts) to outperform benchmarks, it is computationally cheap and outperforms alternative methods even with significantly fewer observations as input. These features make the NowcastPNN model a promising tool for now-casting in epidemiological surveillance of arboviral threats and other domains involving right-truncated data.

## 1 Introduction

Delayed reporting of infectious disease cases, healthcare admissions, and deaths often hinders accurate real-time outbreak analyses, epidemic situation reports, as well as analyses of ongoing or endemic pathogen threats. Indeed, while new diagnoses, healthcare admissions, or deaths occur on well-defined, recorded dates, these data remain widely subject to data entry or data collection delays in modern surveillance systems. Following delayed data entry, real-time reporting systems tend to retrospectively update past cases, healthcare admissions, or death records according to the date on which these occurred. In some systems, however, records may also be added to alternative dates, such as the end of a week or the middle of a month. Characteristically, reporting delays often change rapidly over time—for instance, during the initial stages of novel disease outbreaks, when reporting systems are still being established. They also commonly vary across several factors, such as reporting locations and population strata. These characteristic features mean that traditional statistical methods tend to perform relatively poorly at *now-casting* ; i.e., the estimation of the actual number of infectious disease case counts, healthcare admissions, or deaths that occurred close to real time.

Early examples of nowcasting methodologies included the analysis of delayed HIV diagnosis data (Lawless, 1994), the influenza A/H1N1 outbreak in 2009 in the Netherlands (Donker et al., 2011), and efforts to nowcast the Escherichia coli O104:H4 outbreak in Germany (Höhle and an der Heiden, 2014). In Brazil, Infodengue (Codeco et al., 2018) is a joint effort supported by the Brazilian Ministry of Health, other local health authorities, and research institutions to provide timely information and estimates to decision-makers regarding the number of dengue cases at national, state and municipality levels and the notification delay is corrected using a Bayesian chain-ladder model (Bastos et al., 2019). During the COVID-19 pandemic, nowcasting methods were extended to account for changes in delay distributions over time and across days of the week (Günther et al., 2021). Additionally, auxiliary data sources, such as Google search frequencies, have been utilised for nowcasting influenza outbreaks (Preis and Moat, 2014; Lampos et al., 2015).

To capture complex, non-linear trends in reporting delays more accurately, a range of machine learning (ML) architectures have been proposed, focusing on capturing long-term dependencies and modeling uncertainty. Long Short-Term Memory (LSTM) recurrent neural networks (RNNs) have shown success in time series applications such as precipitation nowcasting (Shi et al., 2015) and macroeconomic prediction (Hopp, 2022), making them well-suited for sequential data.

More recently, attention mechanisms (Vaswani et al., 2017) have enabled deep-learning architectures to selectively focus on specific parts of the input data, thus often substantially improving both efficiency and accuracy in learning tasks in fields such as natural language processing and image recognition (Galassi et al., 2020; Zheng et al., 2017; Qin et al., 2017). Here, we introduce the NowcastPNN model, a novel neural network architecture that builds on attention mechanisms for nowcasting infectious disease case counts in the context of irregular, dynamic reporting delay patterns. The NowcastPNN model further integrates techniques to quantify uncertainty around nowcast point estimates. Probabilistic neural networks (PNNs) (Specht, 1990) extend traditional neural network architectures by estimating distribution parameters instead of single outputs, making them particularly valuable for interpreting infectious disease case counts that are commonly modelled using generative distributions such as the Poisson or Negative Binomial. In addition to capturing generative (aleatoric) uncertainty, we also aim to capture uncertainty in the distribution parameters themselves (epistemic uncertainty) by incorporating Monte Carlo (MC) Dropout (Gal and Ghahramani, 2016) into the NowcastPNN model. Taken together, the proposed architecture outputs generative probabilistic distributions of nowcasted case counts, along with point estimates and margins of error for the distribution parameters themselves, which correspond to the expected case count per time unit and other model parameters such as overdispersion around the expected case count. To the best of our knowledge, this is the first study to evaluate a nowcasting neural network architecture that combines these three features—namely, attention, probabilistic generative outputs, and Monte Carlo dropout—for capturing complex, long-term delay effects and quantifying aleatoric and epistemic uncertainty.

Here, we explore and evaluate the NowcastPNN model for nowcasting daily confirmed dengue cases in the state of São Paulo, Brazil, over six months spanning 2018–2019. The model is trained on surveillance data from 2013–2018 to learn delay patterns. Dengue fever is a mosquito-borne viral infection caused by the dengue virus and transmitted by *Aedes* spp. mosquitoes. Around 5.7 billion people live in areas at risk of dengue infection, with risk being highest in tropical and subtropical regions (Waterman and Gubler, 1989). Brazil, due to its large population, favourable climate, and widespread distribution of *Aedes* spp. mosquitoes, consistently bears a high burden of dengue, frequently accounting for the majority of reported cases in the Americas (Siqueira Jr. et al., 2005; Kraemer et al., 2019). The frequency and severity of outbreaks continues to rise, with a historical record of over 10 million cases in 2024 (World Health Organization, 2025), surpassing the previous annual peak of 1.7 million in 2015 (Dias et al., 2024). Dengue fever has also become more deadly in recent years, with over 1,000 deaths recorded annually since 2015, more than half of which occurred in São Paulo (Andrioli et al., 2020). Additionally, inter-epidemic periods appear to be shortening, further increasing the burden on public health organisations. In this context, nowcasting of dengue case counts has emerged as an important tool to address increasing public health challenges in Brazil through providing near-real-time estimates into the actual confirmed case counts.

This paper is structured as follows. In Section 2, we describe the data obtained from DATASUS, the Brazilian Ministry of Health (Ministério da Saúde). Section 3 presents the methodology, beginning with a definition of the nowcasting task and the evaluation metrics, followed by a description of the proposed NowcastPNN architecture. We also introduce two state-of-the-art models used for benchmarking. Section 4 evaluates the performance of the NowcastPNN method in comparison with the benchmark models. Finally, in Section 5, we summarise the results, discuss their implications, and outline directions for future work.

## 2 Infectious disease case data

The data used in this study were sourced from the Brazilian Ministry of Health (Ministério da Saúde) and the associated DATASUS service, covering the period from 2013 to 2020 for São Paulo (SP), Brazil. Although the dataset includes counts for all states, we specifically focus on SP due to its high number of dengue cases and the presence of the most comprehensive surveillance system, as noted by Andrioli et al. (2020). Each observation contains the date on which symptoms occurred and the date on which the observation was reported in DATASUS, thus providing the information that is needed to assess reporting delay patterns in the nowcasting task. Each case in the dataset was reported with an associated date of occurrence, enabling daily-level analysis and nowcasting. In our setting, at least 99% of cases were reported within a maximum delay of 40 days (Figure 2, bottom left panel).

While it is common to include environmental covariates in dengue case predictions, this paper focuses on adjusting for reporting delays inherent in the surveillance system using only the available confirmed case counts and the day of the week (DOW) on which the report was made, as delays may vary significantly by weekday. The potential for incorporating additional information to enhance estimation accuracy within the NowcastPNN framework is discussed in Section 3.2, and further detailed in Supplementary Section SS1.

## 3 Methods

### 3.1 Nowcasting data triangle

Nowcasting is based on time series data of retrospective case counts, healthcare admission, or death counts that have been retrospectively updated as time progresses and reporting delays become known. We consider regularly incrementing time inputs *s* = 1, … , *t* up to the most recent calendar time *t*, and define *x*_*s*,*d*_ as the number of cases that were measured at time *s* and were reported with a delay of *d* = 0, 1, … , *D* time units. These counts can be calculated either from occurrence and reporting dates associated with each observation, or reconstructed from multiple versions of retrospectively updated time series data. We consider time in units of days. Thus, the final confirmed case count, death or hospital admission at time *t* is defined as

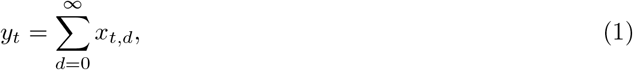

and similarly for all times *s < t*. In practice, reporting delays are limited to a certain delay horizon *D >* 0 such that *x*_*s*,*d*_ = 0 for all *d* ≥ *D*, or such that the vast majority of cases that were measured on day *s* have been reported. Note that *x*_*s*,*d*_ was reported and thus became known on day *s* + *d*. Table 1 illustrates that, close to real time *t*, all reported counts within the delay horizon *t*^*^ ∈ *{t* − *D* + 1, … , *t}* are biased from below because some of the delayed counts *x*_*t*\*,*d*_ are in the future and thus have not yet been observed, as indicated in red. The nowcasting task is to make accurate and reliable predictions *y*_*t*_* for all time inputs within the delay horizon close to real time *t, t*^*^ ∈ *{t* − *D* + 1, … , *t}*.

**Table 1:** Illustration of the reporting triangle at time *t*, such that *D* = 6 and *M* = *t*. Quantities known at time *t* are shown in black, while unknown quantities are shown in red.

| Time \ Delay | $d = 0$ | $d = 1$ | $d = 2$ | $d = 3$ | $d = 4$ | $d = 5$ | Final count |
| --- | --- | --- | --- | --- | --- | --- | --- |
| 1 | $x_{1,0}$ | $x_{1,1}$ | $x_{1,2}$ | $x_{1,3}$ | $x_{1,4}$ | $x_{1,5}$ | $y_1$ |
| 2 | $x_{2,0}$ | $x_{2,1}$ | $x_{2,2}$ | $x_{2,3}$ | $x_{2,4}$ | $x_{2,5}$ | $y_2$ |
| ⋮ | ⋮ | ⋮ | ⋮ | ⋮ | ⋮ | ⋮ | ⋮ |
| $t - 5$ | $x_{t-5,0}$ | $x_{t-5,1}$ | $x_{t-5,2}$ | $x_{t-5,3}$ | $x_{t-5,4}$ | $x_{t-5,5}$ | $y_{t-5}$ |
| $t - 4$ | $x_{t-4,0}$ | $x_{t-4,1}$ | $x_{t-4,2}$ | $x_{t-4,3}$ | $x_{t-4,4}$ | $x_{t-4,5}$ | $y_{t-4}$ |
| $t - 3$ | $x_{t-3,0}$ | $x_{t-3,1}$ | $x_{t-3,2}$ | $x_{t-3,3}$ | $x_{t-3,4}$ | $x_{t-3,5}$ | $y_{t-3}$ |
| $t - 2$ | $x_{t-2,0}$ | $x_{t-2,1}$ | $x_{t-2,2}$ | $x_{t-2,3}$ | $x_{t-2,4}$ | $x_{t-2,5}$ | $y_{t-2}$ |
| $t - 1$ | $x_{t-1,0}$ | $x_{t-1,1}$ | $x_{t-1,2}$ | $x_{t-1,3}$ | $x_{t-1,4}$ | $x_{t-1,5}$ | $y_{t-1}$ |
| $t$ | $x_{t,0}$ | $x_{t,1}$ | $x_{t,2}$ | $x_{t,3}$ | $x_{t,4}$ | $x_{t,5}$ | $y_t$ |

The primary information for the nowcasting prediction task is the structure and regularity in the observed reporting delays *x*_*s*,*d*_ before the delay horizon, *s* = 0, … , *t* − *D*. In addition, partially observed delay counts *x*_*t*\*,*d*_ within the delay horizon (such that *t*^*^ + *d* ≤ *t*) inform if historic patterns in reporting delays still apply, or how they may be changing towards real time. As data are typically less relevant the further they lie in the past, historic records before time *t* − *M* for some non-negative *M* are typically ignored. For these reasons, predictions at time *t*^*^ are based on the so-called “nowcasting data triangle”

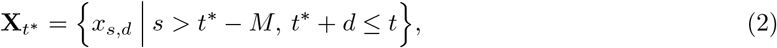

where *t* denotes the current calendar time, *t*^*^ ∈ *{t* − *D* + 1, … , *t}*, and *M* is specified by the user. In our applications, we set *M* = *D* = 40 days, at which point 99% of confirmed dengue cases across the entire observation period had been reported during the study period. Sensitivity analyses reported in the Supplementary Material suggest *M* could be set to values as low as *M* = 10. The counts shown in black in Table 1 illustrate the reporting triangle at the most recent calendar time *t*. To train nowcasting models, we consider time series data before the delay horizon such that the counts in Equation (1) are accurately observed, together with the corresponding nowcasting data triangle and potentially other user-specified features **S**_*s*_,

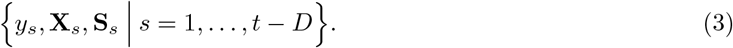

### 3.2 NowcastPNN

We now describe the main components of the NowcastPNN model as summarized in Figure 1. For each time point *t* during training (and similarly for the time points *t*^*^ during prediction), the input data consists of the reporting triangle **X**_*s*_ in Equation (2) in matrix form of dimension *M × D* and additional user-specified features **S**_*s*_, for which we here consider the day of week. As a first step, **X**_*s*_ is passed through an attention block, where the delay horizon *D* is used as the embedding dimension and the individual delay values represent the embedding. The internal dimensions of the attention block are configured to ensure that the output has the same dimensions as the input. To retain information of the original input, we added the output of the attention block to the model input **X**_*s*_ via a residual (or skip) connection following common practice (Vaswani et al., 2017). The resulting matrix is then passed through two convolutional layers to reduce the *M × D* matrix to a feature vector with a single value per past unit of dimension *M* . The day of the week **S**_*s*_ is mapped to a 10-dimensional vector, which is processed by dense layers, and its output is added to the output from the convolutional layers. The combined vector is subsequently passed through two stacked dense layers, gradually reducing its length from *M* to two non-negative values that represent the latent mean and overdispersion parameters *µ*_*s*_, *ϕ*_*s*_ of the Negative Binomial (NB) distribution in our probabilistic architecture. The following sections describe each component of the NowcastPNN model, with further details and numerical experiments provided in the Supplementary Material.

**Figure 1:**
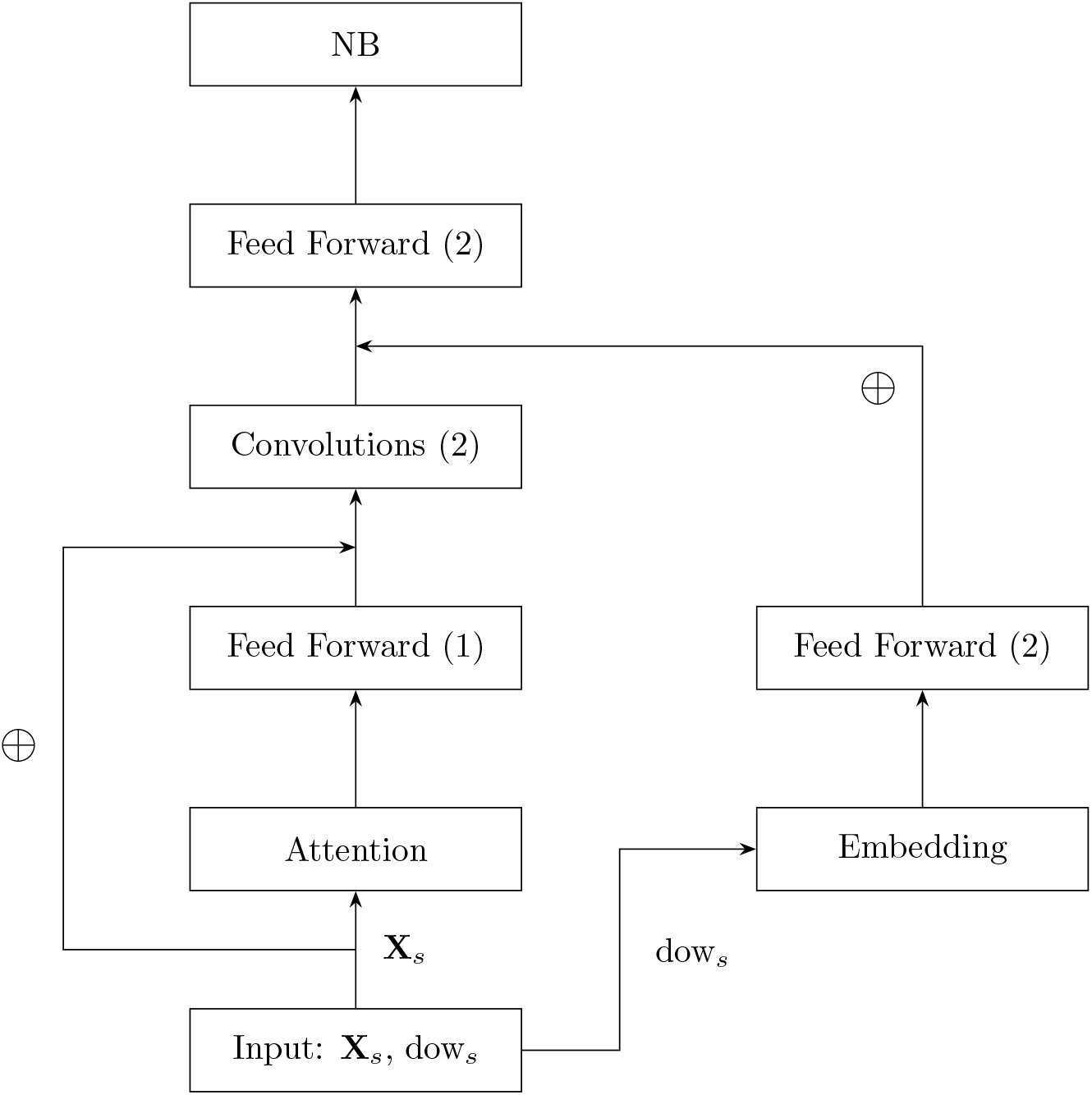
Illustration of the architecture of the NowcastPNN, where the numbers in brackets indicate the number of layers used. All convolutional layers and feed-forward components use the SiLU activation function and batch normalisation, while ⊕ denotes element-wise addition. **X**_*s*_ is the reporting triangle and dow_*s*_ represents the day of the week corresponding to date *s*.

**Figure 2:**
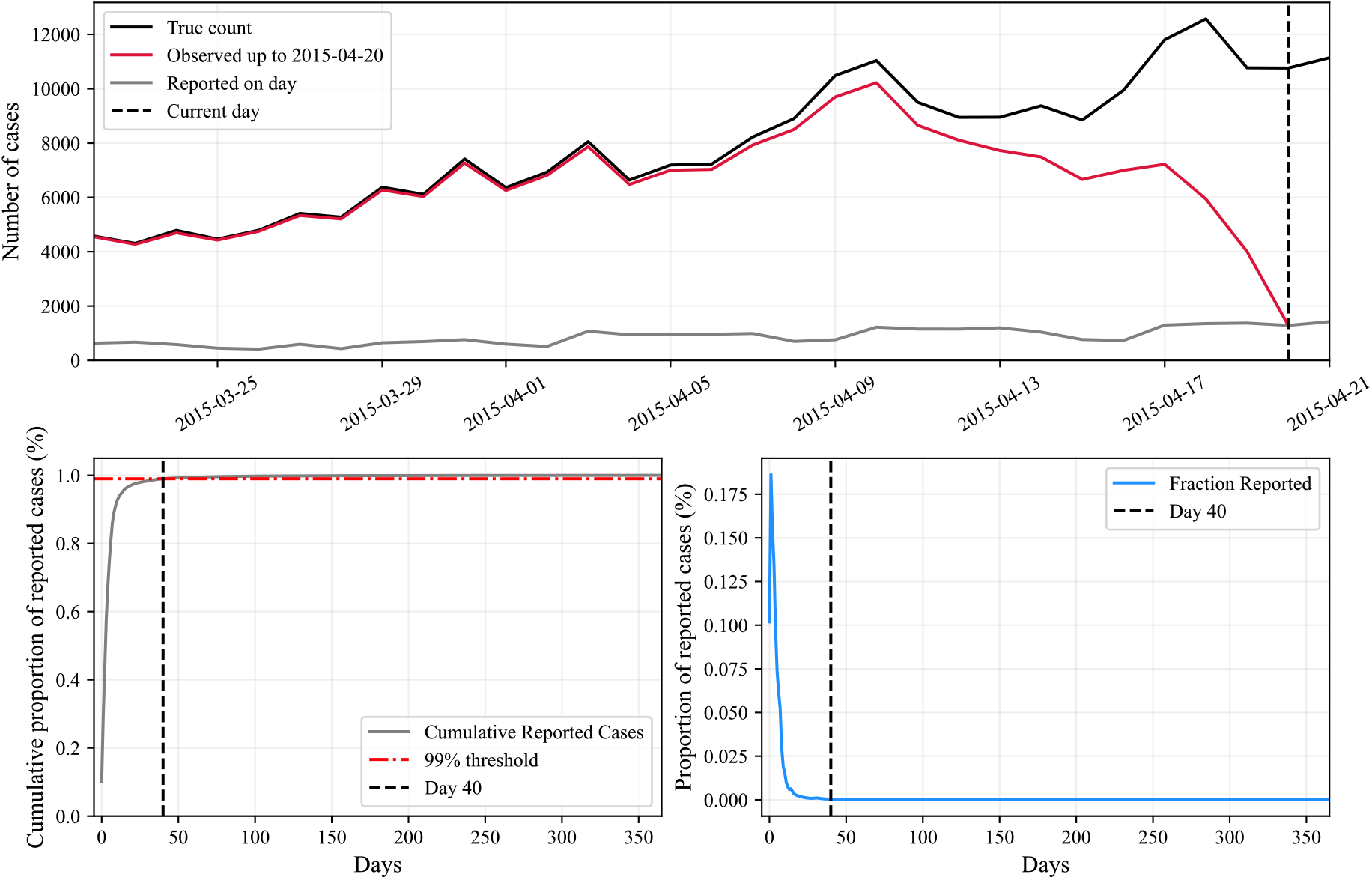
Top panel: Number of dengue cases in São Paulo, Brazil, from March 22, 2015, to April 22, 2015. The red line represents data available in real time as of the forecast date (April 20, 2015, indicated by the vertical dashed line), the black line represents the upward-revised data, and the gray line represents the uncorrected data. Bottom left panel: Cumulative proportion of cases reported with a delay of *d* days (2013–2020). Bottom right panel: Proportion of cases reported with a delay of *d* days (2013–2020).

#### 3.2.1 NowcastPNN probabilistic frontend

Instead of predicting a single case count value for each time input, our PNN frontend returns a probability distribution with input-specific parameters for each time point. This distribution can be used to obtain characteristics such as the (log-)probability, mean, variance, or to generate samples. While non-probabilistic NNs producing point predictions are trained using loss functions, such as the Euclidean (squared error) or cross-entropy loss, PNNs are trained by maximising the log-likelihood of the observed (target) value under the predicted distribution (Specht, 1990). For nowcasting, various statistical models have been proposed (Lawless, 1994; Donker et al., 2011; Bastos et al., 2019; Hawryluk et al., 2021), and here we adopted the Negative Binomial generative model due to its ability to account for overdispersion. In practice, optimisers minimise loss functions and so our PNN frontend target loss is the negative log-likelihood (LL) evaluated at the observed case counts *y*_*s*_ in the training set,

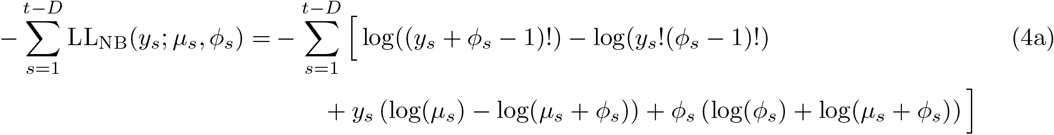

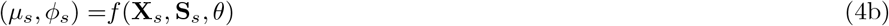

where *µ*_*s*_ and *ϕ*_*s*_ are respectively the distributional Negative Binomial mean and overdispersion parameters for day *s* estimated by the the internal, deterministic layers *f* of the NowcastPNN. The internal layers *f* use as inputs the reporting triangle **X**_*s*_ and other user-specified features **S**_*s*_ for the training points before the delay horizon, *s* = 1, … , *t* − *D*. Note that before the delay horizon, we have that *y*_*s*_ either equals 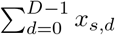 , or is very close to this sum. We refer to all unknown parameters of the NowcastPNN by *θ*, and these parameters are optimised during training as described further below.

In terms of implementation, the Negative Binomial distribution in mean-overdispersion parameterisation is not included in the standard PyTorch distributions library (Paszke et al. (2019), version 2.4.0). We used the open-source implementation available at https://github.com/silaskoemen/NowcastPNN/blob/main/src/NegativeBinomial.py, which also includes an infinite-mixture Poisson-Gamma representation, and replaces the factorial in Equation (4a) with Gamma functions to ensure differentiability for gradient-based optimization.

#### 3.2.2 NowcastPNN attention component

Intuitively, the attention component can be thought of as a mapping that assigns weights to different elements of the input reporting triangle, indicating the importance of each element relative to the others in generating the output. In the attention component, for each time point *s* the input reporting triangle **X**_*s*_ ∈ R^*M ×D*^ is transformed into three matrices, namely the query (**Q**_*s*_), key (**K**_*s*_), and value (**V**_*s*_) matrices, by multiplication with learned weight matrices **W**^Q^, **W**^K^ and 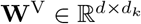. These transformations result in **Q**_*s*_, **K**_*s*_, and **V**_*s*_ of dimensions *n × d*_*k*_, where *d*_*k*_ is the size of the latent space used for the attention mechanism. To preserve the original feature dimension, *d*_*k*_ is often set equal to *d*, which in our case is the maximum delay *D*. The scaled dot-product attention is then computed as

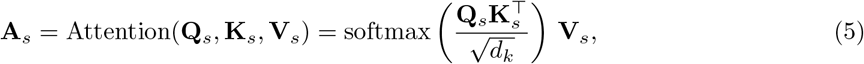

where **Q**_*s*_, **K**_*s*_, **V**_*s*_ ∈ ℝ^*M ×D*^, 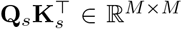 and thus 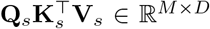 (element-wise softmax and scalar division by 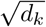 does not change the dimension) is the same dimension as the input **X**_*s*_, such that the output of the attention components can be added element-wise to the input. This process allows the model to selectively focus on the most relevant parts of the input reporting triangles at each step. Note that we compute in Equation (5) pairwise attention scores and so focus is extended to relevant data points several time steps away in either direction, and not restricted to purely sequential patterns as is common in other architectures like e.g., the LSTM layer (Hochreiter and Schmidhuber, 1997).

As is standard practice, the output of the attention layer is processed by a dense layer, followed by an activation function, to allow for non-linear relationships, before being added to the input data to retain information about the original input. All these processing steps can be summarized as the attention block.

#### 3.2.3 NowcastPNN day-of-week embedding

The NowcastPNN architecture enables embedding additional user-specified features into the learning task to allow for greater expressiveness in learning true case counts, possibly including information that is not included in the reporting delay patterns themselves. We investigated and exemplified this with integer-valued day-of-week inputs for each time point *s*, **S**_*s*_ ∈ *{*Monday = 0, … , Sunday = 6*}*. The day-of-week input is converted into a higher-dimensional real-valued vector. As the embedding layer adapts during training, the model learns the optimal position for each day in the embedding space. The embedding layer is followed by two dense layers that further process the vector corresponding to the day of the week, where the output of the last dense layer has a dimension of *M* , ensuring that the length of the vector matches that of the output from the convolutional layers, so they can be added together. While embedding dimension, number of dense layers, and units per dense layer are parameters that can be adapted by the user, an embedding dimension of 10 and dense layers with 20 and then *M* = 40 units with batch normalisation showed great performance.

#### 3.2.4 NowcastPNN feed forward and convolutional layers

As mentioned above, the *M × D* dimensional outputs from the attention layer are then passed into a depth-1 feed-forward dense layer (Gurney, 2018) to create activated attention scores,

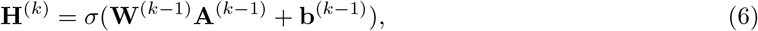

where 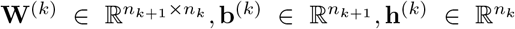 are the weight matrix, bias vector, and post-activations, respectively. *σ*(·) denotes the activation function, or non-linearity, and *n*_*k*_ denotes the number of hidden units in the *k*^th^ layer. Naturally, **h**^(0)^ = **x**, the output of dimension *M × D* from the attention layer, and consequently, *n*_0_ correspond to the length or dimension of the input data *M* . As dense layers operate on vectors, each column of the resulting matrix is processed separately, corresponding to each vector of past counts for each delay value. Denoting **a**_*d*_ as the vector containing the output of the attention layer for column *d* ∈ *{*0, … , *D* − 1*}* along the delay dimension, the outputs for each column are calculated as **h**_*d*_ = *σ*(**Wa**_*d*_ + **b**) and can subsequently be stacked to generate the matrix of activated attention scores **H**_*d*_ = [**h**_1_, … , **h**_*D*−1_]. More generally, the final model predictions are given by ŷ = *σ*_out_(**W**^(*L*)^**h**^(*L*)^ + **b**^(*L*)^) for a total of *L* hidden layers.

The *M × D* dimensional outputs from the Feed-Forward block for each time point *t* are then passed into a depth-2 convolutional neural network component in order to reduce the activated attention scores **H**_*t*_ to an *M* dimensional feature vector through two one-dimensional convolutional layers with a filter of length 7 to capture weekly patterns.

For a 1D convolutional layer, each observation **X**_*t*_ is represented as an *M × D* matrix, where *M* is the length (or temporal dimension) and *D* ≡ *C*_in_ is the number of input channels. The output for an observation **X**_*t*_ and output channel *j* is given by

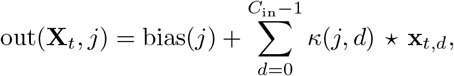

where **x**_*t*,*d*_ denotes the *d*-th input channel of **X**_*t*_ (i.e., the *d*-th column of size *M* ), *κ*(*j, d*) are the kernel weights for output channel *j* and input channel *d*, and ⋆ denotes the cross-correlation operation—also known as the sliding dot product (Pinaya et al., 2020).

All trainable parameters *θ* then consist of the kernel weights and biases of the two convolutional layers, weights and biases of all dense layers, as well as the learnable parameters of the batch normalisation layers and the attention layer. For the model without the weekday embedding, |*θ*| = 13, 849, whereas the additional dense layers, embedding layer, and batch normalisation layer used in the processing of the day of the week bring up the total number of learnable parameters to |*θ*_dow_| = 14, 959.

#### 3.2.5 NowcastPNN uncertainty quantification

Quantifying uncertainty is crucial for nowcasting tasks in infectious disease epidemiology, as in many cases nowcasting outputs serve as inputs into downstream analytics such as estimating effective reproduction numbers, into which nowcasting uncertainty needs to be incorporated for credible epidemic trend analyses. Although fully Bayesian neural networks have been proposed (Kononenko, 1989; Tishby et al., 1989), they often underperform or entail significant computational costs, making them impractical for widespread use. Even with approximation techniques such as variational inference (Goan and Fookes, 2020; Blundell et al., 2015), the computational burden remains a substantial challenge. Here, we adopted a heuristic alternative, MC dropout (Gal and Ghahramani, 2016). While dropout layers have been used successfully at training time to generate more robust model predictions, their use during inference leads to an ensemble of model architectures, each with slightly different architectures (due to the dropout layers zeroing out certain connections). Specifically, we used dropout layers before the two intermediate dense layers in the final dense part of the NowcastPNN model (and not before the final dense layer outputting the distributional parameters (*λ*_*s*_, *ϕ*_*s*_)). These dropout layers independently drop connections between layers, specified by layer-specific dropout probabilities *p*_*k*_ following a Bernoulli distribution, where inference can be done in parallel, to create an ensemble of different NowcastPNN parameters (*µ*_*s*_, *ϕ*_*s*_, *θ*)^*r*^ for *r* = 1, … , *R*. For inference, we choose *R* = 200 so we can build all empirical uncertainty intervals from individual draws. We heuristically interpret the uncertainty in this ensemble as epistemic uncertainty in the true model parameters, which is separate to the generative (or aleatoric) uncertainty captured in the Negative Binomial PNN component in Equation (4). We explored potential dropout probabilities numerically, and found the range [0.1, 0.3] performed best in order to generate valid uncertainty intervals while also benefitting from more generalizable model predictions due to using the dropout layers during model training (Supplementary Table ST2).

### 3.3 NowcastPNN training

To train the NowcastPNN and specify all tuning parameters, we split the data before the delay horizon 75%–25% into a training data and test set. We considered two approaches, the first involved a random split of the data before the delay horizon, which we refer to as the “random test set,” while the second approach split the data such that the most recent 25% of observations were used for testing, which we refer to as the “recent test set.” The training data was again split 75%–25% into training and validation set to monitor the NowcastPNN’s performance on an independent validation set. The optimal probabilities for the dropout layers before the first and second dense layers were found to be *p*_1_ = 0.15 and *p*_2_ = 0.1 when evaluated on the randomly split test set, while *p*_1_ = 0.3 and *p*_2_ = 0.1 were optimal for evaluation on the most recent observations as test set. Recall that these dropout probabilities both influence model training (by encouraging more generalizable model predictions), as well as performance on test sets by affecting the width of uncertainty intervals.

Throughout, we used PyTorch’s built-in Adam optimiser (Kingma, 2014) to optimize the free model parameters *θ* against the PNN negative log Negative Binomial loss in Equation (4). Optimization was done for up to 200 epochs, with early stopping applied if no improvement on the validation set was observed for 30 consecutive epochs. After training, the model weights were set to those achieving the best performance on the validation set, to ensure optimal out-of-sample performance and to prevent overfitting. To achieve this, model weights were saved and automatically updated whenever improved validation performance was recorded, such that the final saved weights correspond to the optimal model.

#### 3.3.1 NowcastPNN predictions

Provided a fitted NowcastPNN model, we numerically generated nowcast predictions *y*_*t*_* at time inputs *t*^*^ according to the heuristic MC dropout approximation (Gal and Ghahramani, 2016) to the log Bayesian posterior predictive density

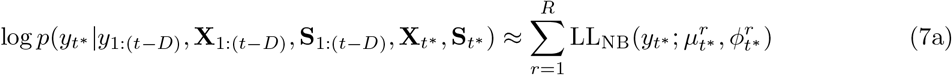

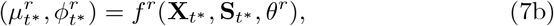

where the time inputs are in the delay horizon close to real time *t, t*^*^ = *t* − *D* + 1, … , *t*, and *f* ^*r*^ are the internal layers of the NowcastPNN subject to the *r*-th node dropout and *θ*^*r*^ are the corresponding optimised latent parameters under our training loss in Equation (4) on training data up to time *t* − *D*. Thus, unlike other dropout applications (Srivastava et al., 2014), we retained epistemic dropout uncertainty for prediction.

### 3.4 Evaluation metrics

We assessed model performance through the interval score (IS) (Gneiting and Raftery, 2007), the weighted interval score (WIS) Bracher et al. (2021) and prediction interval coverage accuracy (PICA), which focus on accounting for the extent of predictive uncertainty in assessing model performance. For these scores, we first constructed central (1 − *α*) *×* 100% posterior prediction intervals (PIs) from the trained model for specific time inputs *t*^*^, with *α* ∈ (0, 1). For each level *α*, we define the interval bounds as *L*_*α*_ = *F*^−1^(*α/*2) and *U*_*α*_ = *F*^−1^(1 − (*α/*2)), where *F* is the empirical predictive cumulative distribution function. The set of significance levels used was *A* = *{*0.05, 0.10, 0.25, 0.5, 0.75, 0.9, 0.95*}*.

PICA focuses on evaluating the absolute difference between expected and actual coverages across a set of specified levels *α* ∈ *A*. For a specific level *α*, we define

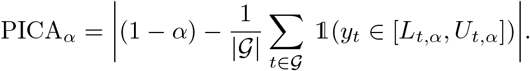

In the following, PICA (without a subscript) denotes the average over all levels, given by 1*/*|𝒜| ∑_*α*∈*A*_ PICA_*α*_. Note all scores are negatively oriented, meaning lower values reflect better performance.

The IS is computed using

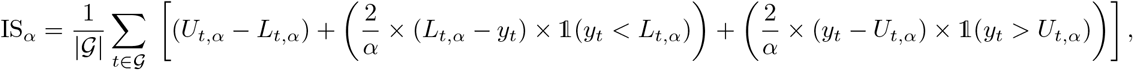

where 1 denotes the indicator function, *t* are the indices in the hold-out test set *G*, and *L*_*t*,*α*_ and *U*_*t*,*α*_ are the estimated lower and upper bounds of observation *y*_*t*_ at the coverage level *α*. Similarly to before, IS (without a subscript) will refer to the average over all levels, given by 1*/*|𝒜| ∑_*α*∈*A*_ IS_*α*_. Note that the interval score is defined as the sum of three components that penalise *spread, underprediction*, and *overprediction*, and we used this decomposition to gain a more detailed understanding of model predictions.

The WIS score is based on the IS, however, it weighs the individual components by the corresponding levels, i.e.,

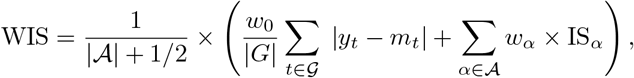

where *w*_*α*_ = *α/*2, with *w*_0_ = 1*/*2, and *m*_*t*_ is the predictive median. The WIS is a discrete approximation of the Continuously Ranked Probability Score (CRPS) (Hersbach, 2000) and can be thought of as a generalized version of the mean absolute error (MAE) for distributions or interval predictions. The WIS is also a proper scoring rule, meaning it has the lowest expected score if the predicted distribution of the target is equal to the true distribution (Höhle and an der Heiden, 2014), and thus encourages honest predictions.

### 3.5 Benchmarking

We benchmarked the NowcastPNN against two state-of-the-art nowcasting tools that were among the best-performing models across various metrics in a comparative study by Wolffram et al. (2023), Epinowcast (Abbott et al., 2021) and the RIVM model (van de Kassteele et al., 2019).

Epinowcast employs a semi-parametric Bayesian model that accounts for factors such as reporting delays and missing data, and is implemented in the Stan probabilistic programming language (Carpenter et al., 2017). It supports various probability distributions for case occurrences within groups and automatically adjusts for assumptions about maximum delays. Epinowcast assumes that all delay counts for at least one data point are observed to estimate the reporting delay distribution. In our application, this implies that at least *D* = 40 days of recorded history are required as input to nowcast the present day.

Epinowcast has been used, for example, to nowcast COVID-19 in Germany, Mpox outbreaks in the UK, and influenza incidences in the US.

The RIVM model (van de Kassteele et al., 2019) is another Bayesian method using bivariate penalised spline smoothing of the reported cases by time of symptoms onset and reporting delay to produce the nowcast. The method assumes that the underlying reporting intensity surface is smooth and incorporates prior information about the reporting process as constraints. Moreover, the surface is assumed to be unimodal in the reporting delay dimension and nearly zero at the delay horizon *D*. The RIVM model has been applied, for example, to nowcast COVID-19 hospitalisation incidences in Germany.

## 4 Results

### 4.1 Improved uncertainty quantification with NowcastPNN

For the time period from 2013-01-01 to 2020-12-31, we retrieved in total 3,557,329 confirmed dengue case records in the state of São Paulo (SP), Brazil, with available date of onset and date of reporting from the DATASUS service of the Brazilian Ministry of Health. Figure 2 (top panel) illustrates the typical reporting delays in confirmed dengue case records by symptom onset date. Over the entire time period, at least 99% of the cases were reported within a delay horizon 40 days, as shown in Figure 2 (bottom left panel).

Figure 3 shows the same-day nowcasts of confirmed dengue case counts in São Paulo by symptom onset date for the time period December 2018 to July 2019, using the Epinowcast, the RIVM, and NowcastPNN models. This time period was part of our recent test set, and all models were trained on data from 2013-02-09 to 2018-12-12. Same-day nowcasting refers to the scenario where the model, at each time point, only has access to data reported up to that moment and forms predictions accordingly. A visual comparison of same-day nowcasts over the entire test set period is provided in Figure SF5 (Supplementary Material). Overall, we found the Epinowcast and RIVM models tended to generate more frequently posterior median same-day nowcasts that deviated significantly both upwards and downwards from the true counts compared to the NowcastPNN. Additionally, the prediction intervals of the Epinowcast and RIVM models were very wide, reducing their informativeness. For instance, when the true daily count reached approximately 8,000 cases, the Epinowcast’s 95% posterior prediction intervals exceeded 17,000, and the RIVM model’s 95% posterior prediction intervals exceeded 13,000, whereas the NowcastPNN’s 95% posterior prediction intervals exceeded only 10,000 cases.

**Figure 3:**
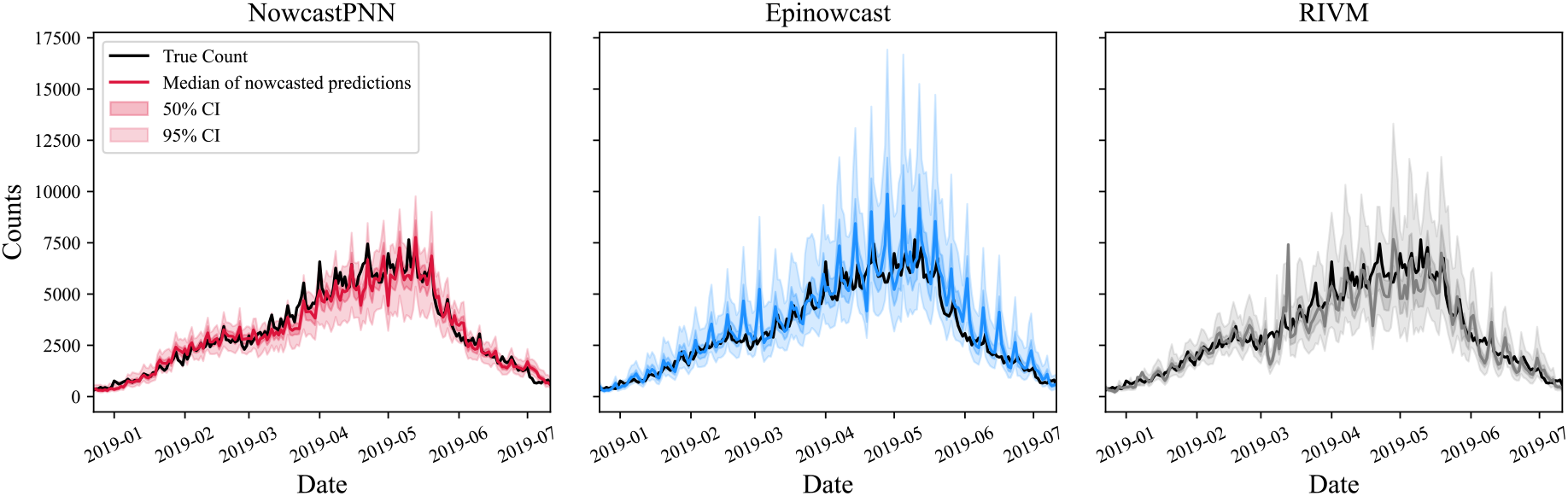
Comparison of same-day nowcasts produced by all models from December 2018 to July 2019, as part of the *recent test set*.

We evaluated model performance with a particular focus on uncertainty quantification systematically with the interval score (IS), the weighted interval score (WIS), and the prediction interval coverage accuracy (PICA) using either randomly chosen time point as test set (random test set), or the most recent time points from 2018-12-13 to 2020-11-22 as test set (recent test set) (see Section 3.3).

These metrics focus on assessing whether the proportion of actual dengue case counts that fell within the posterior prediction intervals—once calendar time had moved beyond the delay horizon—matched the expected levels; in other words, whether the posterior predictive coverages aligned closely with the nominal coverages. Figure 4 illustrate the relationship between the actual and expected coverages of the *α* = 50% and 95% prediction intervals, along with the PICA Score, which quantifies the average absolute deviation between the posterior predictive and expected coverages across all levels of *α*. These results are shown for the random and recent test sets, respectively. While all models achieved similar PICA scores, the Epinowcast and NowcastPNN models aligned more closely with the desired levels than the RIVM model on both the random test set and the recent test set. This was particularly the case on the recent test set, where the PICA score of the RIVM model was more than twice as high as those achieved by Epinowcast and NowcastPNN.

**Figure 4:**
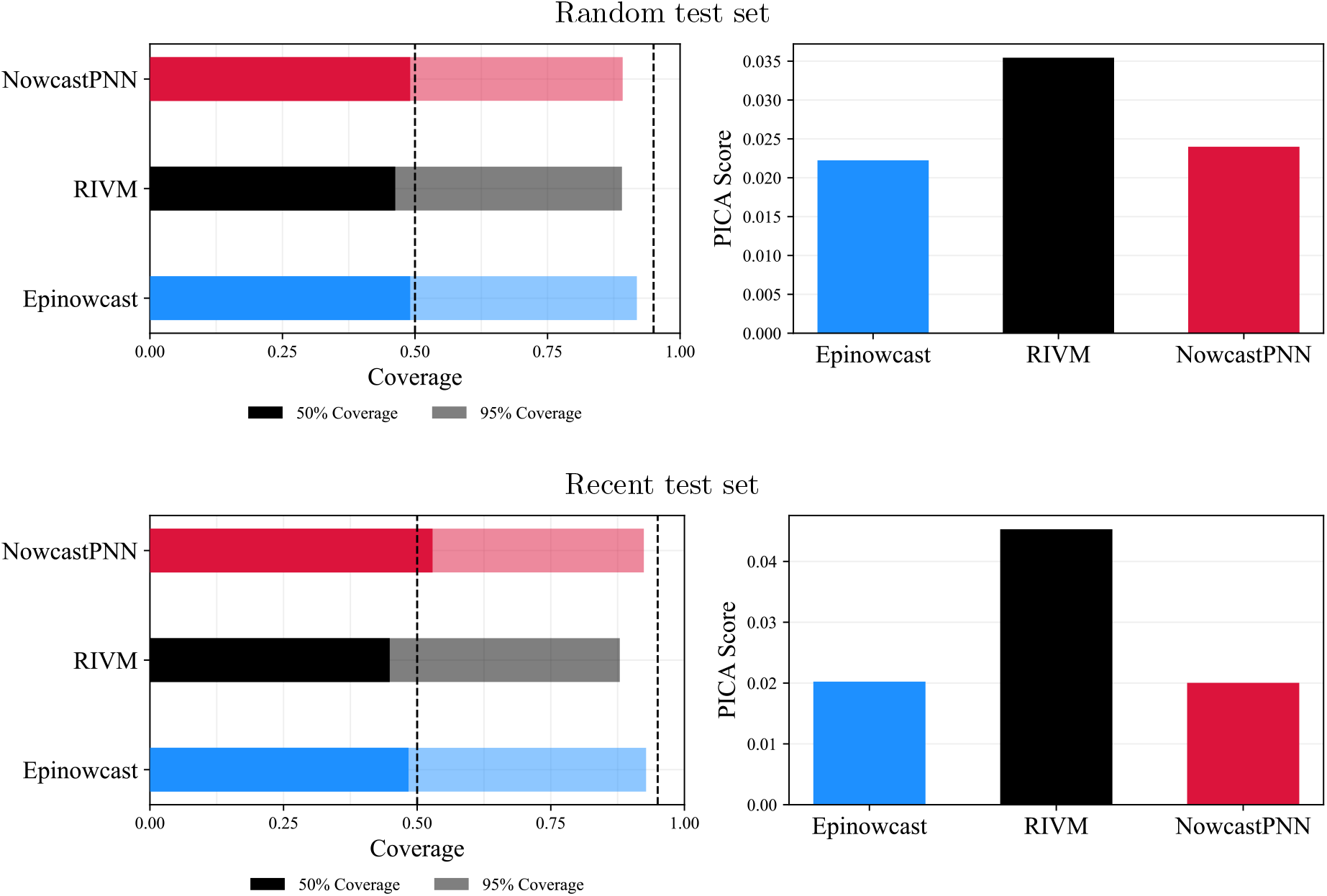
Left column: Empirical coverage at 50% and 95% levels. Right column: PICA score for all models. Top panels: Evaluated on the *random test set*. Bottom panels: Evaluated on the *recent test set*.

Beyond coverage, the Interval Scores (IS) and Weighted Interval Scores (WIS) penalise the width of uncertainty intervals and extent of over- and underprediction. As shown in Figures 5, the NowcastPNN outperformed the other two models on the random and recent test sets in IS and WIS. The RIVM model tended to underpredict actual case counts more frequently than the other two methods, while the Epinowcast model tended to overpredict actual case counts more frequently than the other two methods. While both over- and underprediction are undesirable, underprediction can be particularly problematic for infectious disease management and policy making. In this particular context, we found that Epinowcast was associated with the lowest underprediction error and outperformed the other two methods on the random and recent test sets.

**Figure 5:**
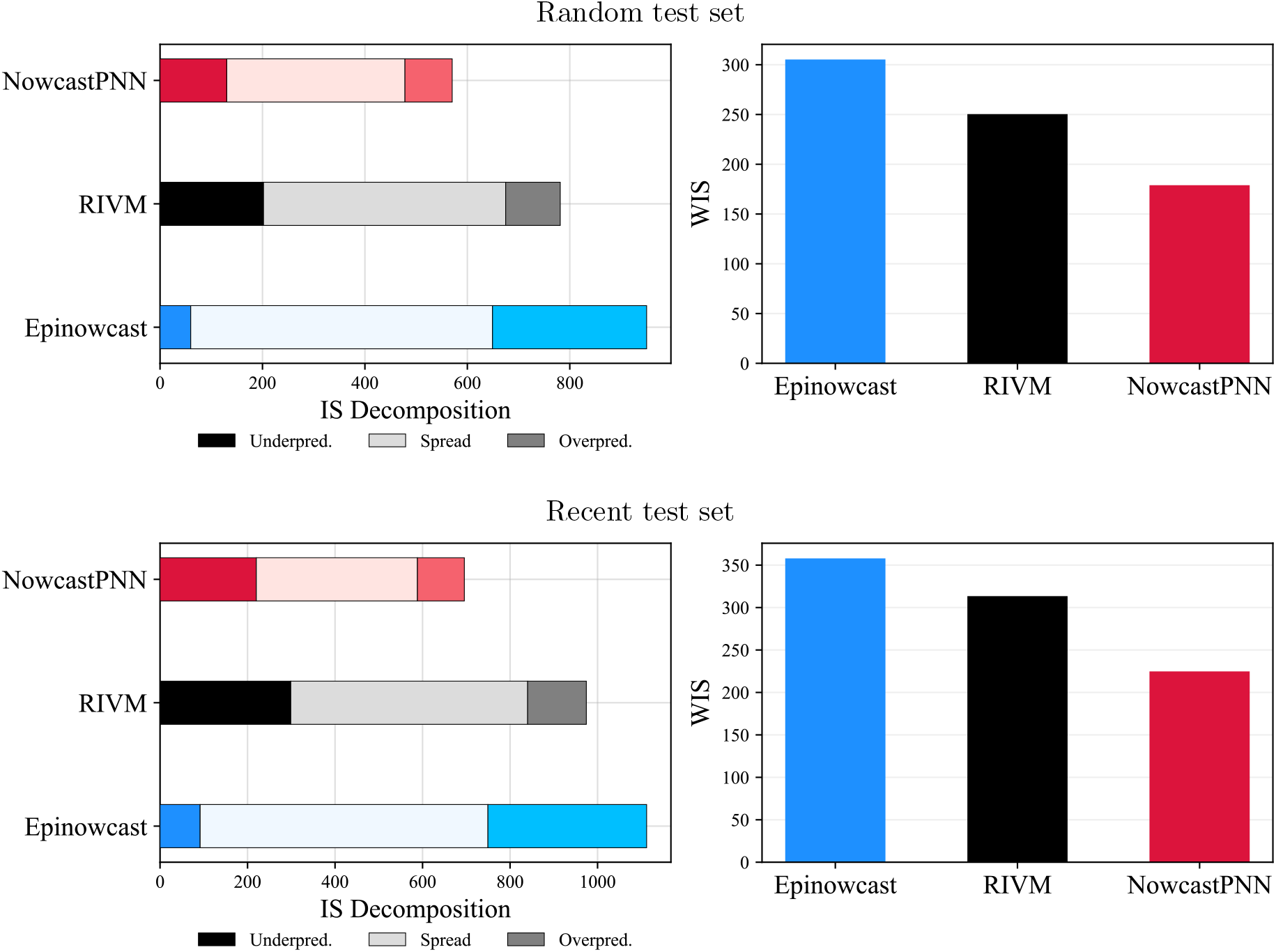
Left column: Interval Score (IS). Right column: Weighted IS (WIS) for all models. Top panels: Evaluated on the *random test set*. Bottom panels: Evaluated on the *recent test set*.

Further evaluation details are provided in Supplementary Table ST1. Also, it is important to note that the scores achieved on the recent versus random test sets cannot be directly compared, as the magnitudes of incidences and interepidemic periods differ. Scores should only be compared between models within the same test set. Overall, these results demonstrate that by combining deep learning architectures with a probabilistic distributional frontend, it is possible to obtain state-of-the-art or better uncertainty quantification around infectious disease case count nowcasts, including same-day nowcasts by day of symptom onset.

### 4.2 NowcastPNN requires separate training data

While the Epinowcast and RIVM models estimate the distributional parameters of the delay counts locally for each observation, the NowcastPNN model requires separate training data to learn the model weights used for later inference. In terms of computational runtimes, it took under 10 minutes on a MacBook Air with M1 Chip (with GPU training acceleration using the Metal Performance Shaders (MPS) backend) to train the NowcastPNN on the full training data set of 2133 daily records (75% of data), with the version without day-of-week embeddings training slightly faster (an average of roughly 8 minutes).

In many other contexts, especially emerging diseases, less training data may be available and we therefore evaluated the impact of providing fewer training data points on nowcasting performance. We found that on the recent test set, 1,500 training data points (corresponding to 4.1 years of training data) were required for the NowcastPNN to achieve comparable performance to Epinowcast and the RIVM model in terms of PICA, IS and WIS. On the random test set, 1,000 data points (corresponding to 2.74 years of training data) were required. Further details are provided in the Supplementary Section SS9.2. Moreover, Supplementary Section SS9.1 investigates whether giving the Epinowcast and RIVM models more data for estimation (i.e. more past observations) leads to better predictions, but the NowcastPNN still outperforms both models even with 80 past observations as input.

### 4.3 Faster prediction with NowcastPNN

As previously discussed, the RIVM and Epinowcast models operate fundamentally differently from the NowcastPNN. In the case of RIVM and Epinowcast, each prediction step involves re-estimating distributional parameters based on the corresponding reporting triangle, after which individual nowcasts are generated by sampling from the fitted distributions. In contrast, the NowcastPNN follows a machine learning approach: it learns optimal parameters (i.e., model weights) during a separate training phase. Once trained, predictions with NowcastPNN consist solely of computing the distributional parameters of the assumed Negative Binomial distribution based on the fixed, learned weights, removing the need for additional fitting and significantly speeding up the prediction process. Table 2 shows the mean prediction times for an individual nowcast from all models. Both versions of the NowcastPNN model achieve significantly faster prediction times, with the slower DOW variant achieving more than 280 times faster predictions than Epinowcast and almost 16 times faster than the RIVM model.

**Table 2:** Average prediction times (in seconds) for all models. Timings correspond to the mean prediction time for the first 20 observations of the *recent test set*, with standard deviations shown in parentheses.

| NowcastPNN | NowcastPNN (DOW) | EpiNowcast | RIVM |
| --- | --- | --- | --- |
| 0.142 (0.010) | 0.182 (0.014) | 51.661 (7.104) | 2.900 (0.973) |

Note that these estimates of the inference time for the NowcastPNN model are conservative, as sampling can be further accelerated through parallelisation, and NowcastPNN allows for batched predictions across multiple observations simultaneously. For example, generating predictions for the entire test set of 711 observations took only 12.82 seconds for the model without and 13.87 seconds for the model with DOW embeddings. Further details on computing times and the computational setup are provided in the Supplementary Material.

### 4.4 Uncertainty quantification does not necessarily improve as calendar time progresses

As calendar time progresses from day *t*− 1 to *t*, additional information in the nowcasting data triangle **X**_*s*_ becomes available, allowing for an improved same-day nowcast for day *t* − 1, as well as for all other time points within the updated delay horizon *t*^*^ ∈ *{t* − *D* + 1, … , *t}*. This motivated us to evaluate nowcasting prediction performance at time points *t*^*^ by the number of days *s* = 0, 1, … lagging behind the current calendar time *t, t*^*^ = *t* − *s*. In general, for all nowcasting models, posterior median estimates tended to be closer to the actual values as additional information became progressively available with increasing lag days *s*, as shown in Figure 6 (and Supplementary Figures SF6–SF8). However, as expected, credible intervals also became narrower with the addition of more information. For this reason, it does not necessarily follow that uncertainty quantification becomes more trustworthy with increasing lag days *s*. We found, in fact, that the coverage of all three models worsened as more future observations were included (Figure 6). This effect was particularly pronounced for the RIVM model, where, for example, the 95% prediction interval with 13 additional observations captured fewer than 50% of actual cases.

**Figure 6:**
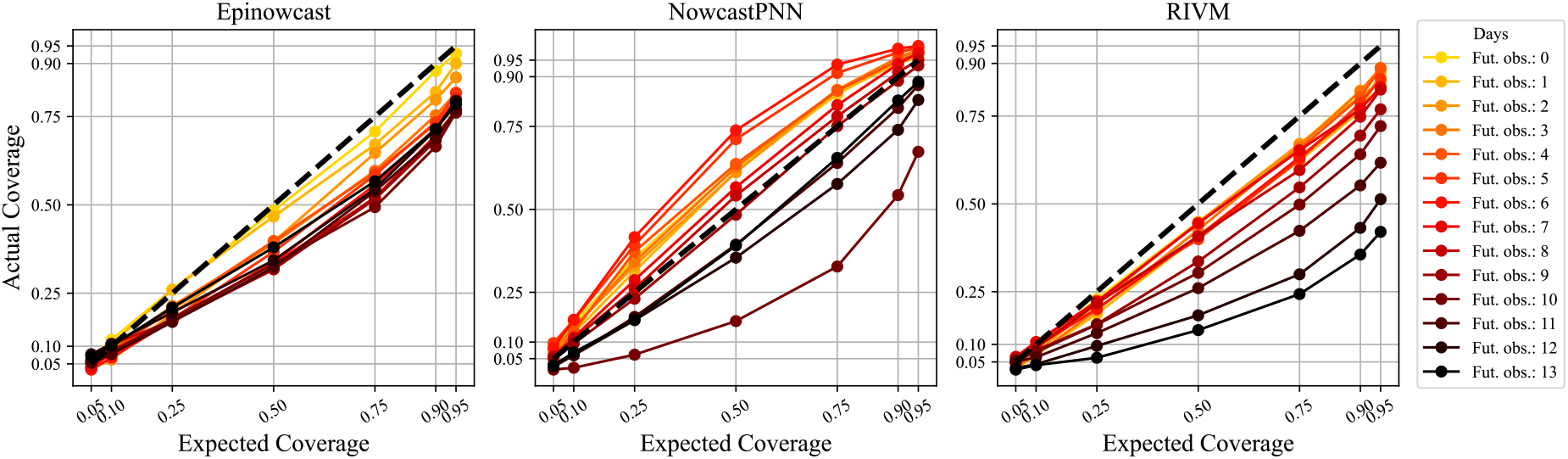
Comparison of expected and actual coverages for all models as more future observations become available. The dashed identity curve denotes perfect alignment. Evaluation was performed on the *recent test set*.

We next considered the IS and WIS error scores as a compromise between posterior median prediction accuracy and uncertainty quantification trustworthiness. Figure 7 shows that IS and WIS generally tended to decline as additional information became available for Epinowcast and NowcastPNN, though not for the RIVM model. For Epinowcast, the decline in nowcasting error was more rapid than for NowcastPNN; however, NowcastPNN achieved lower IS and WIS scores than both Epinowcast and the RIVM model for up to three days lag behind the current calendar time.

**Figure 7:**
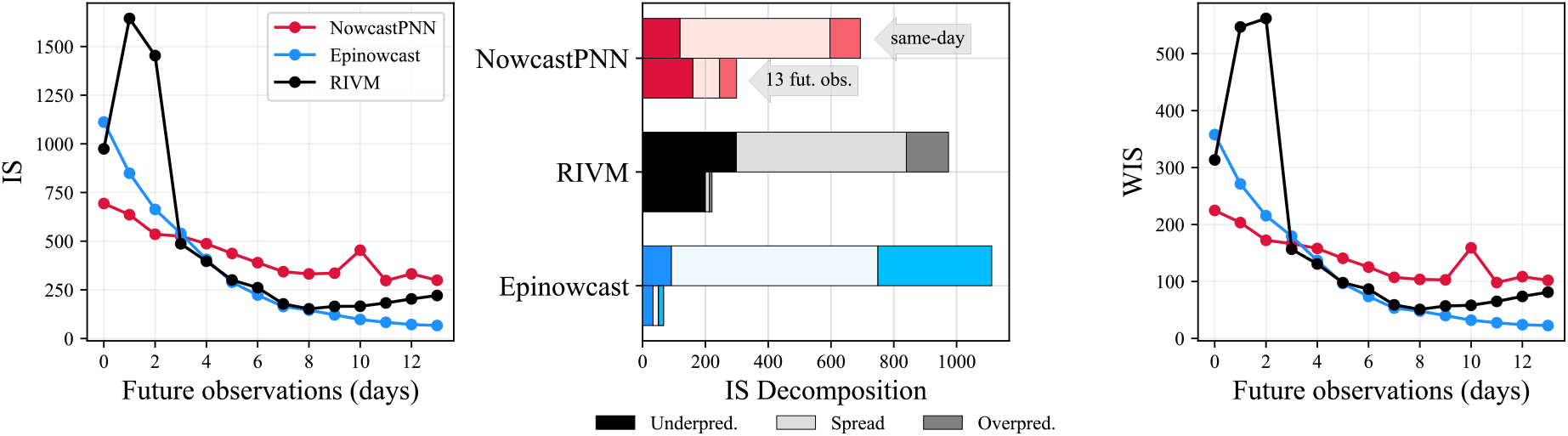
Left: Interval Scores (IS) as a function of the number of future observations. Middle: IS decomposition for same-day nowcast and after 13 days of additional observations. Right: Weighted IS (WIS) as a function of the number of future observations. Evaluation was performed on the *recent test set*.

## 5 Discussion

In this paper, we introduce the NowcastPNN, a probabilistic neural network-based model for now-casting symptomatic, confirmed cases of febrile dengue virus infections by date of symptom onset in the state of São Paulo, Brazil, between 2013 and 2020. We demonstrate that it is possible to develop an attention-based neural network architecture that enables orders-of-magnitude faster training and—critically—generates trustworthy error magnitudes around same-day nowcasts, capturing and disentangling both epistemic and aleatoric uncertainty in case counts. Due to the expressiveness of our deep learning architecture, large training sets spanning 3–4 years of past observations are required to achieve state-of-the-art predictive performance.

On the São Paulo dengue case data, the NowcastPNN model outperformed two benchmark models, the Epinowcast and the RIVM models, representing a substantial improvement in nowcasting prediction performance with uncertainty quantification. Specifically, we found that the NowcastPNN architecture was able to generate prediction uncertainty ranges that were in good agreement with theoretically expected coverage levels, and had IS and WIS scores 25–40% lower than those of the two benchmark models. Moreover, our model was highly computationally efficient, with training completed in minutes on a personal computer and predictions generated within seconds, making it well-suited for real-time epidemiological surveillance, particularly if predictions for many different locations are required quickly. For dengue specifically, this could also facilitate nowcasting of particular genotypes or major lineages (Hill et al., 2024), which would aid characterisation of their potential to cause outbreaks in different settings. Although the default settings of the model provided good results, its flexible architecture can support additional fine-tuning at multiple levels, allowing extensions to a variety of other settings, including the nowcasting of febrile dengue cases across municipalities in Brazil (Lopes and Bastos, 2025) and the nowcasting of other infectious disease case counts, healthcare admissions, or deaths. For example, this flexibility could be of immediate public health significance in the face of ongoing outbreaks caused by the chikungunya and Oropouche viruses in Brazil (Scachetti et al., 2025).

Our findings need to be considered in the context of several limitations. First, nowcasting methods generally only aim to adjust for reporting delays, assuming that all cases, healthcare admissions or deaths are fully reported after a certain delay horizon. The predictions generated by nowcasting methodologies therefore do not capture unreported, unattended symptomatic, misattributed, or asymptomatic cases. Second, the model requires relatively large amounts of historical data for training, which is limited for newly emerging diseases or infectious disease outbreaks in previously unmonitored regions. A possi-ble solution may involve training on data from diseases with the same transmission vector (e.g., Zika and chikungunya) or different geographical areas such as those available in the OpenDengue database (Clarke et al., 2024) or the dengue global surveillance dashboard from the World Health Organization (World Health Organization, 2025). Third, we only included day-of-week features as additional input features into the NowcastPNN, while many more external covariates such as climate patterns, population movements, or case counts from neighbouring regions could potentially further enhance predictive accuracy (Harish et al., 2024). Fourth, as shown in Amaral et al. (2024), combining forecasts from multiple models—an approach known as *ensemble nowcasting* —or applying post-processing techniques to the nowcasts can enhance predictive performance. Thus, although NowcastPNN outperformed two benchmark models, integrating its predictions with those of other models could further refine estimates and improve robustness. Finally, as with most machine learning models, if the prediction data differ substantially from the training data, the model performance may decline. For example, in the event of a new outbreak significantly larger in magnitude than those seen in the training data, the model may underestimate case counts. Future research could explore ways to augment the training data to address this limitation.

In conclusion, these characteristics establish NowcastPNN as a valuable tool for infectious disease modelling and public health decision-making. By generating accurate real-time nowcasts, it becomes particularly useful for outbreak response, allowing health authorities to anticipate case surges and allocate resources efficiently. Rapid training and inference time enable public health officials to produce a large number of nowcasts rapidly, even in low-resource settings. In addition, the probabilistic framework provides uncertainty estimates, which are essential for risk assessment and policy planning. With a flexible and tunable architecture, the model can be easily adapted to ensure precise estimates with valid confidence intervals, regardless of case counts, transmission dynamics, or other factors. Moreover, its applicability extends beyond dengue fever, making it suitable for a wide range of nowcasting scenarios.

## Acknowledgments

This study was supported by the Wellcome Trust–Digital Technology Development Award (226075/Z/22/Z) (NRF); the Medical Research Council and São Paulo Research Foundation (MR/S0195/1 and FAPESP 18/14389-0) (NRF); the World Health Organization–Temasek Foundation Collaboration Framework Grant (NRF); the International Pathogen Surveillance Network Catalytic Grant Fund (NRF); and the Engineering and Physical Sciences Research Council (EPSRC) (OR and NRF). NRF also acknowledges funding from the MRC Centre for Global Infectious Disease Analysis (MR/X020258/1), funded by the UK Medical Research Council (MRC). This UK funded award is carried out in the frame of the Global Health EDCTP3 Joint Undertaking. LSB acknowledges funding from the Fundação Carlos Chagas Filho de Amparo `a Pesquisa do Estado do Rio de Janeiro (E-26/204.098/2024) and the National Council for Scientific and Technological Development (310530/2021-0).

## Data availability

The daily case counts of dengue fever in São Paulo from 2013 through 2020, indexed by symptom onset and reporting date, are available as an open-access dataset from Zenodo (https://zenodo.org/records/15292499) under the CC-BY-4.0 license.

## Code

The NowcastPNN model was implemented in PyTorch (Paszke et al., 2019) and is available as open-source software at https://github.com/silaskoemen/NowcastPNN.

